# Mapping research priorities related to maternal and infant nutritional health in the context of climate change in Pakistan: A mixed methods study

**DOI:** 10.1101/2025.11.22.25340614

**Authors:** Sadiq Bhanbhro, Zahid Memon, Wardah Ahmed, Fizza Ansar, Gulfishan Tahir, Nadia Agha, Sally-Fowler Davis, Hora Soltani

## Abstract

Climate change poses significant threats to health, food security, and nutrition, particularly in Pakistan, which is vulnerable due to its limited resources and preparedness. Maternal and infant health targets remain largely unmet due to several factors. However, there is a lack of a relevant people, institutions, and place-informed agenda from those most affected, such as rural women, that guides research, policy, and practice. We employed a mixed-methods explanatory design, engaging key individuals and groups, and integrating an online survey with workshops to gather both quantitative and qualitative perspectives. Overall, 127 people participated in the study: 88 in the survey, 24 in the priority-setting workshop, and 15 in the focus group session. The participants included community members, doctors, lady health workers, nurses, nutritionists, climate change experts, advocates, and researchers. The survey data were analysed using the STATA statistical package, and the qualitative data were analysed thematically. Priority ranking exercises revealed consensus across surveys and workshops on nutritional (“lack of awareness of proper nutrition”) and maternal (“dehydration, weakness, and exhaustion”) challenges. However, infant health priorities diverged; surveys ranked “heat exposure and low birth weight” highest, while workshop participants prioritised “heat waves and infant hospital admissions”. Furthermore, half of the participants ranked developing climate-resilient nutritional interventions and addressing the nutritional needs of pregnant women in a changing climate as equally critical. Focus group narratives consistently highlighted how climate-induced events, such as extreme heat, unseasonal rainfall, and recurrent floods, disrupted agricultural cycles, displaced families, and cascaded into effects on families, particularly pregnant and breastfeeding women. There is an urgent need to address nutritional awareness, strengthen climate-resilient nutritional strategies, and provide targeted support for pregnant women. Adaptation and mitigation strategies are urgently needed and should be central to policy responses, alongside the impacts of climate extremes.

## Introduction

Climate change has emerged as one of the most pressing global challenges, posing an imminent and multifaceted threat to public health and well-being. Its effects, including extreme weather events and natural disasters, exacerbate health risks, disrupt food systems, and increase global nutritional insecurity [1]. These impacts are disproportionately severe in low- and middle-income countries (LMICs), such as Pakistan, where limited adaptive capacity hinders resilience against climate-induced “shocks” [2, 3, 4]. Pakistan is introducing mitigation strategies for heat stress, floods, droughts and seawater intrusion [5]. There is a need to analyse projections of climate change for the area in question, Sindh [6], so that the consequences, including those for nutrition and food security, can be estimated.

Rural communities and women are among the most marginalised populations because of challenges related to limited financial and household independence, and often bear the brunt of climate-related adversities [7]. For women, particularly during pregnancy and childbirth, vulnerability increases due to increased physiological and healthcare needs [8]. Effective reproductive health interventions, spanning family planning to prenatal and antenatal care, have been shown to improve maternal and child health outcomes significantly [9]. Despite their proven efficacy, Pakistan falls short of achieving maternal health targets outlined in the Sustainable Development Goals (SDGs) [10].

An increasing body of evidence reports an escalating threat from heat on the rate of pre-term births [11], particularly among those of lower socioeconomic status. The consideration of complications due to climate change, shocks, and longer-term effects such as air pollution has been neglected [12]. Health advice includes adequate hydration, limiting outdoor activities in excess heat, and reducing burning of fossil fuels in the home [13], with nutrition offering an opportunity to mitigate other risks through good general health.

A critical barrier to progress in this area is the absence of a robust, relevant, and community-informed research agenda. In Pakistan, there is a notable gap in research on the intersections of maternal and infant health with climate change-induced weather events and their associated disasters. Moreover, the lack of inclusive research prioritisation processes involving service users, care providers, and policy makers further limits the development and implementation of evidence-based interventions [9].

To address this gap, this mixed-methods study aimed to identify priorities and offer recommendations for research, policy, and practice. The outcomes of this project offer valuable insights for shaping Pakistan’s research agenda, promoting multi-sectoral collaboration, and enhancing maternal and infant health outcomes related to nutrition and climate change.

### Study Objectives

The objectives of this study were threefold. First, we developed an online survey informed by a review of the existing literature and project team discussions to identify research priorities related to maternal health and the impacts of climate change. We then conducted this survey with a wide range of relevant health workers and professionals. Second, we engaged academic, policy, practice, and community members through a priority-setting workshop and focus group session to consolidate, rank, and map the research priorities identified in the survey. Third, to produce a comprehensive list of research priorities and finalise it based on responses from diverse participants, including community members, rural women and men, researchers, healthcare professionals, and policymakers, who participated in the study and workshops.

## Methods

The participants’ recruitment process and the data collection were completed between 02 February 2024 and 15 August 2024, after obtaining required ethics approval from both institutions. This section outlines the ethics statement and the theoretical framework that informed the study, including the research design, settings, participants, data collection tools, data analysis, and the process of obtaining ethical approval.

### Ethics Statement

Before data collection, ethics approval was obtained from the Aga Khan University Ethical Review Committee (ERC-2023-9128-27163) on 25 November 2023 and from the Sheffield Hallam University Research Ethics Committee (ER59568368) on 9 January 2024. All participants provided informed written consent. No minor participants were included in the study. Confidentiality and anonymity were ensured throughout the study. For the online survey, participants indicated consent by selecting the “agree to participate” option after reading the participant information sheet and consent form. Written informed consent was obtained in person prior to participation in the focus group.

### Theoretical framework

This study was informed by a blended theoretical approach drawing on Eco-Social Theory [14] and Intersectionality Theory [15–17]. Eco-social theory emphasises the links between social structures and environmental factors that are embodied in biology, and this epistemology provides a lens for developing data-collection tools that recognise the problems of inequity and susceptibility to disease created by exposure to environmental conditions [14]. Surveys and design-priorities workshops, engage stakeholders in examining how social, biological, and environmental factors jointly shape maternal and infant nutritional health within the broader context of climate change-linked risks. The priority-setting exercises conducted with stakeholders and communities based on previous relevant work (Co-author HS) included the development of Global Midwifery Research Priorities [18] and the establishment of maternal and perinatal health research priorities beyond 2015 [19]. The adopted frameworks informed the integrative analysis, which allowed the project to trace the pathways through which environmental degradation and food insecurity interact with structural determinants, such as poverty, gender norms, and access to healthcare.

Intersectionality Theory complements socio-ecological theory by foregrounding how overlapping challenges, including gender, class, and rurality, shape differential experiences of vulnerability and resilience. Together, these frameworks enabled a multi-level systemic analysis that links micro-, meso-, and macro-level structural conditions with individual and community-level experiences, helping identify context-specific research priorities that reflect both environmental challenges and social inequities.

### Study design

We adopted a sequential explanatory mixed methods design, underpinned by a pragmatist paradigm [20, 21]. Quantitative survey data were first collected to identify broad patterns and priorities related to maternal and infant nutritional health in the context of climate change. These survey results then informed qualitative data collection through a priority setting workshop and focus groups, which provided in-depth insights into lived experiences and local contextual nuances. The pragmatist approach enabled methodological flexibility, allowing the integration of generalisable trends with rich, narrative understandings to inform actionable research priorities. The use of multiple methods ensures a comprehensive and people-informed understanding, that seeks to identify contextually relevant research priorities, and creates actionable recommendations and collaborative strategies to inform research, policy, and practice for improving maternal and infant health outcomes [22] in changing climatic conditions in Pakistan. The mixed-methods study was aligned with the Good Reporting of Mixed Methods Study (GRAMMS) checklist (Supplementary File-1GRAMMS) [23].

### Study setting

The study was conducted from April to May 2024 in Sindh province of Pakistan. Respondents from across Sindh province participated in the survey, and workshops and a focus group were held at Aga Khan University in Karachi and Mother and Child Health Research and Training Centre in Matiari District. Following the 2022 mega floods, the region experienced drought and seawater intrusion into farmland. Climate vulnerability in the region has exacerbated social and economic marginalisation of many communities [5].

### Data collection

#### Online survey

An online survey was developed using Qualtrics (Provo, UT, 2018) from March to April 2024. The semi-structured questionnaire was based on the scoping review of available and relevant literature (Supplementary File-2 Questionnaire). The cross-sectional survey aimed to capture the perspectives of a wider range of health care professionals working across Sindh, Pakistan, at the nexus of climate change, nutrition, and maternal and infant health. Participants were invited to complete the survey via mass email distribution, personal email invitation, and WhatsApp from the authors. In addition to multiple-choice questions, the survey included a few open-ended questions to capture the respondents’ qualitative perspectives.

The survey was pretested with 15 participants, and any errors and issues were corrected, such as for section “Impact of Extreme Heat/Extreme Weather Events on maternal and infant health and nutrition”, the broad response option on breastfeeding was split into two specific options for clarity, while rating scale instructions for section 7 on “Research priorities” were simplified for better readability. The redundant open-ended prompt in section 2, “Awareness and concerns,” was removed to reduce respondent burden. The survey was conducted in English. No sample size target was determined a priori; the researchers aimed to obtain the most relevant sample within the specified time. Respondents involved in the online survey included doctors, nurses, midwives, nutritionists, researchers, climate change experts, policymakers, and public and non-profit organisations working on maternal health, nutrition, and climate risks.

#### Priority setting workshop

A half-day priority-setting workshop was held at the Aga Khan University Karachi campus as part of this study. The workshop included researchers, midwives, nurses, doctors, nutritionists, students, government officials, NGO representatives, civil society actors, and experts on the study theme, including maternal and infant health, nutrition, and climate change. The purpose was to validate, expand, rank, and discuss the contextual nuances of the research priorities identified during the study’s survey phase. The same priority ranking questions from the survey were used to ensure consistency and comparability. Prior to the ranking exercise and facilitated discussions, local academic and policy experts were invited to contextualise the issues by presenting evidence on maternal nutrition and climate-related risks specific to the region. This process enabled critical engagement with diverse knowledge systems, ensuring that identified research priorities reflected both community needs and structural realities.

#### Focus group session

A focus group discussion was conducted in Matiari District, Sindh, to explore community perceptions of the impacts of climate change on maternal and infant nutrition and health. Participants were purposively selected from rural areas of Matiari based on their experiences with climate-induced extreme weather events, including heatwaves and floods. The purposive sampling aimed to identify information-rich cases with lived experiences relevant to the study’s focus. The group included both men and women from the community, ensuring gendered perspectives on climate-related vulnerabilities. The session was conducted in person in the Sindhi language and facilitated by SB, ZM, and FA, with HS present to provide additional prompts, which were translated into Sindhi by the facilitators. In consultation with the Pakistan-based research team, an interview guide was developed in advance in English and translated into Sindhi and Urdu. The session was audio-recorded, transcribed verbatim, and translated into English by a bilingual research associate with expertise in transcription and translation.

#### Participants

In total, 127 individuals participated in the study across its three parts: the survey (n = 88, *see* Table 1), the priority-setting workshop (n = 24) (Supplementary File-3 Participant Demographics) and the focus group session (n = 15, *see* Table 6). Participants included community members, doctors, lady health workers, nurses, nutritionists, climate change experts, advocates, and researchers.

**Table 1.** Demographic characteristics of online survey participants (N=88)

| Variables | Frequency (percentage) |
| --- | --- |
| **Age (years)** |  |
| 18-28 | 19(21.59) |
| 29-39 | 36(40.91) |
| 40-65 | 33(37.50) |
| **Gender** |  |
| Male | 31(35.23) |
| Female | 57(64.77) |
| **Education** |  |
| Secondary School (grade 8th) | 1(1.14) |
| High School (grade 12<sup>th</sup>) | 3(3.41) |
| Bachelor's Degree | 13(14.77) |
| Master's Degree | 59(67.05) |
| Doctoral Degree | 10(11.36) |
| Other | 2(2.27) |

#### Data Analysis

Qualitative data from open-ended survey responses and the focus group session were analysed thematically using the Braun and Clarke approach [24]. The analysis followed a two-stage iterative process. SB, ZM, WA, NA, and FA collaborated on initial coding and theme development in the first stage. In the second stage, an in-person workshop was held at Sheffield Hallam University, bringing together UK- and Pakistan-based team members, including SB, HS, GT, ZM, and WA, to further refine the coding framework and validate emerging themes. Quantitative survey data were analysed using STATA.

### Results and Findings Quantitative Results Online Survey

The online survey was distributed via social media, and N=88 respondents completed it.

#### Demographics

Table 1 below presents Section 1 demographics of the respondents, including distributions by age group, gender, and education level. The age distribution shows that the majority of respondents (41%; n=36) belong to the 29-39 years age group. Regarding gender, 35% (n=31) of participants were male and 65% (n=57) were female. Regarding education levels, 67% of participants held a master’s degree (n=59).

#### Awareness and concerns

section 2 of the questionnaire comprised questions addressing whether the respondents were aware of and concerned about the impact of climate change on maternal and infant nutritional health (Figure 1). Among the total respondents (N = 88), 78 expressed concerns about the impact, while the remaining 10 indicated no concern. Of these, a majority were either extremely (39.74%, n = 31) or very concerned (47.44%, n = 37). Only a smaller % of respondents were moderately or somewhat concerned (7.68%, n=6) in each category. Respondents who expressed concern were asked to include their input in the provided text boxes in the questionnaire.

**Figure 1.**
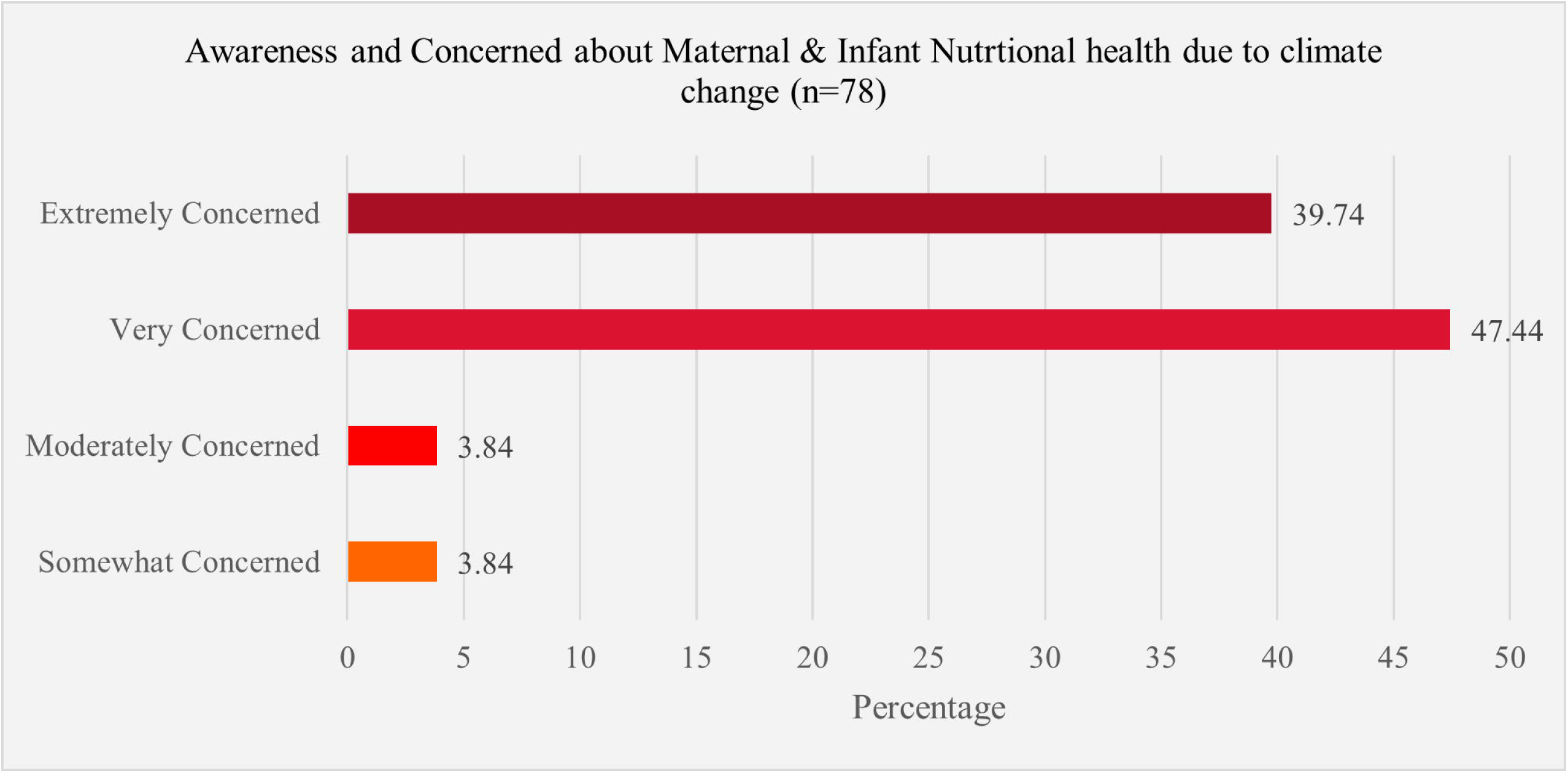
Awareness and Concern about Maternal & Infant Nutritional Health due to Climate Change.

#### Prioritisation of nutritional challenges

The respondents were asked to rank the nutritional challenges pregnant women and infants face in Pakistan in order of priority. The results, summarised from the responses of n=72 participants, are in Table 2. Each challenge was ranked from 1 (highest priority) to 5 (lowest priority), and the percentage distribution for each ranking was shown. Table 2 indicates that “Lack of awareness of proper nutrition” and “Limited availability and accessibility of food” are perceived as the top nutritional challenges for pregnant women and infants in Pakistan. “Limited access to healthcare/nutritional consultation” is also a significant concern across different priority levels. While “Food contamination and safety” and “Climate-related food insecurity” are important, they are generally viewed as lower priorities than the other challenges.

**Table 2.** Prioritisation of Nutritional Challenges faced by pregnant women and Infants.

| **Nutritional Challenges n=72** | **1<sup>st</sup>** | **2<sup>nd</sup>** | **3<sup>rd</sup>** | **4<sup>th</sup>** | **5<sup>th</sup>** |
| --- | --- | --- | --- | --- | --- |
|  | **n (%)** | **n (%)** | **n (%)** | **n (%)** | **n (%)** |
| **Lack of awareness about proper nutrition** | 22(30.56) | 12(16.67) | 16(22.22) | 21(29.17) | 1(1.39) |
| **Limited availability and accessibility of food during climate change-related disasters** | 19(26.39) | 13(18.06) | 15(20.83) | 7(9.72) | 18(25) |
| **Limited access to healthcare/ nutritional consultation during pregnancy** | 12(16.67) | 25(34.72) | 17(23.61) | 7(9.72) | 11(15.28) |
| **Food contamination and safety** | 14(19.44) | 13(18.06) | 13(18.06) | 17(23.61) | 15(20.83) |
| **Climate-related food insecurity** | 5(6.94) | 9(12.5) | 11(15.28) | 20(27.78) | 27(37.5) |

#### Climate change impact on maternal health

The survey included a section focused on understanding the impact of climate change on maternal and infant health. Table 3 presents the percentage distribution of maternal health challenges faced by pregnant women in Pakistan, based on responses from 60 participants. Each challenge was again ranked separately from 1 (highest priority) to 5 (lowest priority).

**Table 3.** Prioritisation of the Climate change impact on maternal health.

| Maternal Health Outcomes | 1<sup>st</sup> | 2<sup>nd</sup> | 3<sup>rd</sup> | 4<sup>th</sup> | 5<sup>th</sup> |
| --- | --- | --- | --- | --- | --- |
|  | n (%) | n (%) | n (%) | n (%) | n (%) |
| Dehydration, weakness, and exhaustion | 25(43.1) | 18(31.03) | 8(13.79) | 5(8.62) | 2(3.45) |
| Access to food | 18(31.03) | 6(10.34) | 6(10.34) | 15(25.86) | 13(22.41) |
| Maternal Mental Health, such as maternal depression, maternal anxiety, worsened mood, etc. | 10(17.24) | 12(20.69) | 12(20.69) | 8(13.79) | 16(27.59) |
| Foetal Development | 4(6.9) | 13(22.41) | 18(31.03) | 13(22.41) | 10(17.24) |
| Breastfeeding or access to safe and affordable alternatives to breast milk | 1(1.72) | 9(15.52) | 14(24.14) | 17(29.31) | 17(29.31) |

The results indicate that “Dehydration, weakness, and exhaustion” and “Access to food” were the most prioritised concerns among respondents regarding maternal challenges. Meanwhile, “Foetal Development” was highlighted as a significant concern, ranking third among participants. “Breastfeeding or access to safe and affordable alternatives to breast milk” was identified as their fourth priority, whereas “Maternal Mental Health, including concerns such as depression, and anxiety” was seen as less prioritised among the respondents.

#### Climate change impact on infant health

Table 4 below presents the percentage distribution of the priority rankings of infant health outcomes related to extreme heat conditions in Pakistan, based on responses from 56 participants. Each outcome, as above, was ranked separately from 1 (highest priority) to 5 (lowest priority). “Increased heat exposure and low birth weight” and “Heat-related infant death” were both seen among the most prioritised concerns. For responses regarding “Extreme heat conditions increase the risks of stillbirth”, it was ranked third. “Heat-related infant death”, “Heat waves increase stress in neonates”, and “Heat waves increase hospital admissions of infants” were relatively prioritised as least concerns, ranking fourth or fifth among the respondents.

**Table 4.**
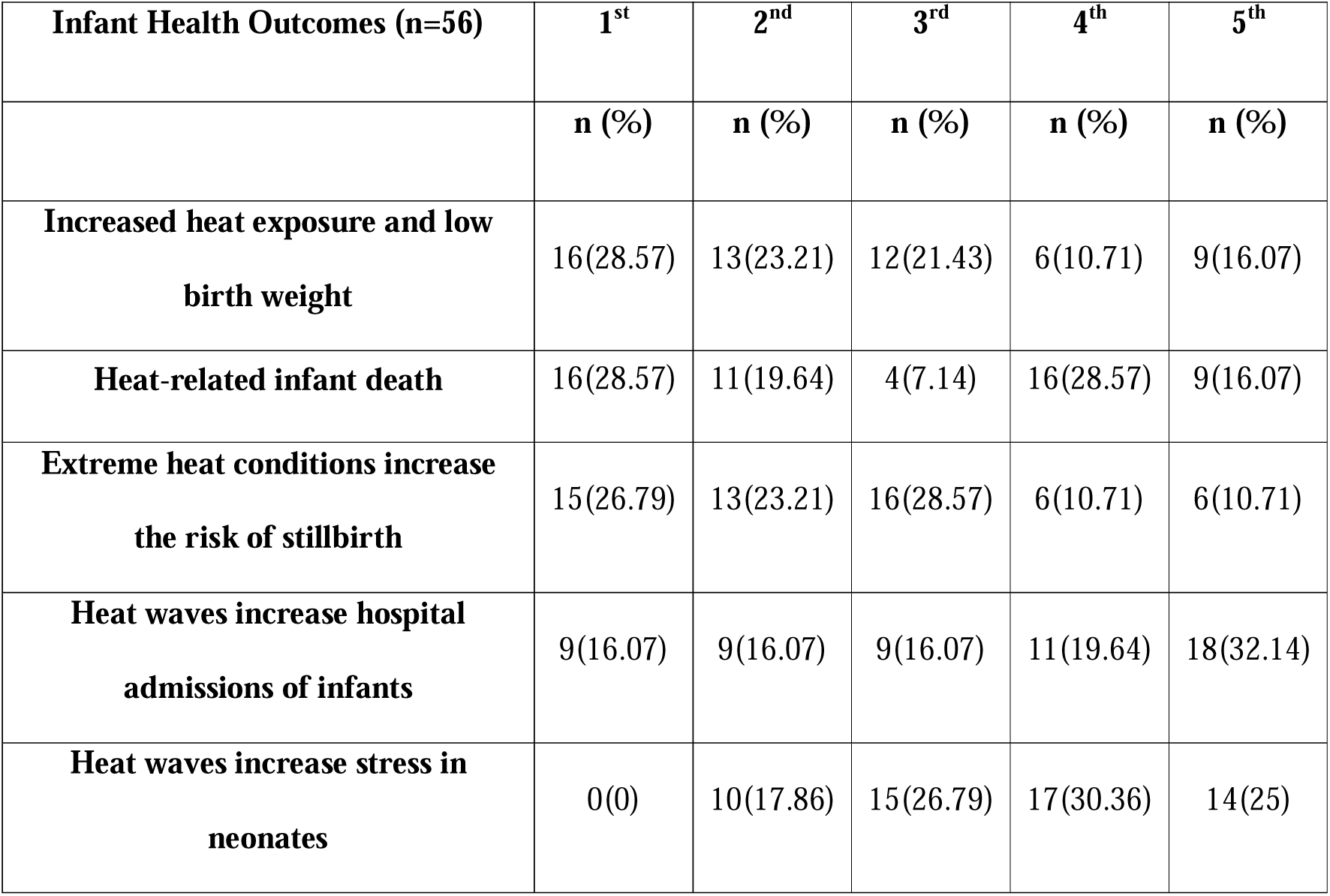
Prioritisation of Climate change impact on infant health.

| **Infant Health Outcomes (n=56)** | **1<sup>st</sup>** | **2<sup>nd</sup>** | **3<sup>rd</sup>** | **4<sup>th</sup>** | **5<sup>th</sup>** |
| --- | --- | --- | --- | --- | --- |
|  | **n (%)** | **n (%)** | **n (%)** | **n (%)** | **n (%)** |
| **Increased heat exposure and low birth weight** | 16(28.57) | 13(23.21) | 12(21.43) | 6(10.71) | 9(16.07) |
| **Heat-related infant death** | 16(28.57) | 11(19.64) | 4(7.14) | 16(28.57) | 9(16.07) |
| **Extreme heat conditions increase the risk of stillbirth** | 15(26.79) | 13(23.21) | 16(28.57) | 6(10.71) | 6(10.71) |
| **Heat waves increase hospital admissions of infants** | 9(16.07) | 9(16.07) | 9(16.07) | 11(19.64) | 18(32.14) |
| **Heat waves increase stress in neonates** | 0(0) | 10(17.86) | 15(26.79) | 17(30.36) | 14(25) |

### Priority-setting workshop

A total of 24 participants participated in the workshop activities, divided into four groups.

#### Priority ranking of challenges

Each group was then asked to discuss and rank the challenges related to the impact of maternal and infant nutrition on climate change to ensure that voices were heard and there would be no facilitator influence. The challenges from the online survey and workshop are tabulated in Table 5 for comparison.

**Table 5.** Priority ranking of challenges related to nutritional, maternal and infant nutritional health impact by climate change.

| **Challenges** | **Ranking in an Online Survey (n=78)** | **Ranking in the priority workshop (n=24)** |
| --- | --- | --- |
| **Nutritional Challenges** |  |  |
| Lack of awareness of proper nutrition | 1<sup>st</sup> | 1<sup>st</sup> |
| Limited availability and accessibility of food during climate change-related disasters | 2<sup>nd</sup> | 3<sup>rd</sup> |
| Limited access to healthcare/nutritional consultation during pregnancy | 3<sup>rd</sup> | 2<sup>nd</sup> |
| Food contamination and safety | 4<sup>th</sup> | 4<sup>th</sup> |
| Climate-related food insecurity | 5<sup>th</sup> | 5<sup>th</sup> |
| **Maternal challenges** |  |  |
| Dehydration, weakness, and exhaustion | 1<sup>st</sup> | 1<sup>st</sup> |
| Access to food | 2<sup>nd</sup> | 4<sup>th</sup> |
| Foetal Development | 3<sup>rd</sup> | 2<sup>nd</sup> |
| Breastfeeding or access to safe and affordable alternatives to breast milk | 4<sup>th</sup> | 5<sup>th</sup> |
| Maternal Mental Health: maternal depression, maternal anxiety, and worsened mood. | 5<sup>th</sup> | 3<sup>rd</sup> |
| **Infant challenges** |  |  |
| Increased heat exposure and low birth weight | 1<sup>st</sup> | 5<sup>th</sup> |
| Heat-related infant death | 2<sup>nd</sup> | 2<sup>nd</sup> |
| Extreme heat conditions increase the risk of stillbirth | 3<sup>rd</sup> | 4<sup>th</sup> |
| Heat waves increase stress in neonates | 4<sup>th</sup> | 3<sup>rd</sup> |
| Heat waves increase hospital admissions of infants | 5<sup>th</sup> | 1<sup>st</sup> |

#### Prioritised research areas

Participants were asked to rate the importance of various research areas related to maternal and infant health in the context of climate change. The scale ranged from 1 (Not Important) to 5 (Extremely Important), and the research areas addressed key challenges, including food availability, nutritional needs, food contamination, climate-resilient interventions, program effectiveness, maternal empowerment, and best practices for reducing maternal malnutrition. A significant proportion of responses rated the above-mentioned research areas as either very important or extremely important, as shown in Figure 2 below:

**Figure 2.**
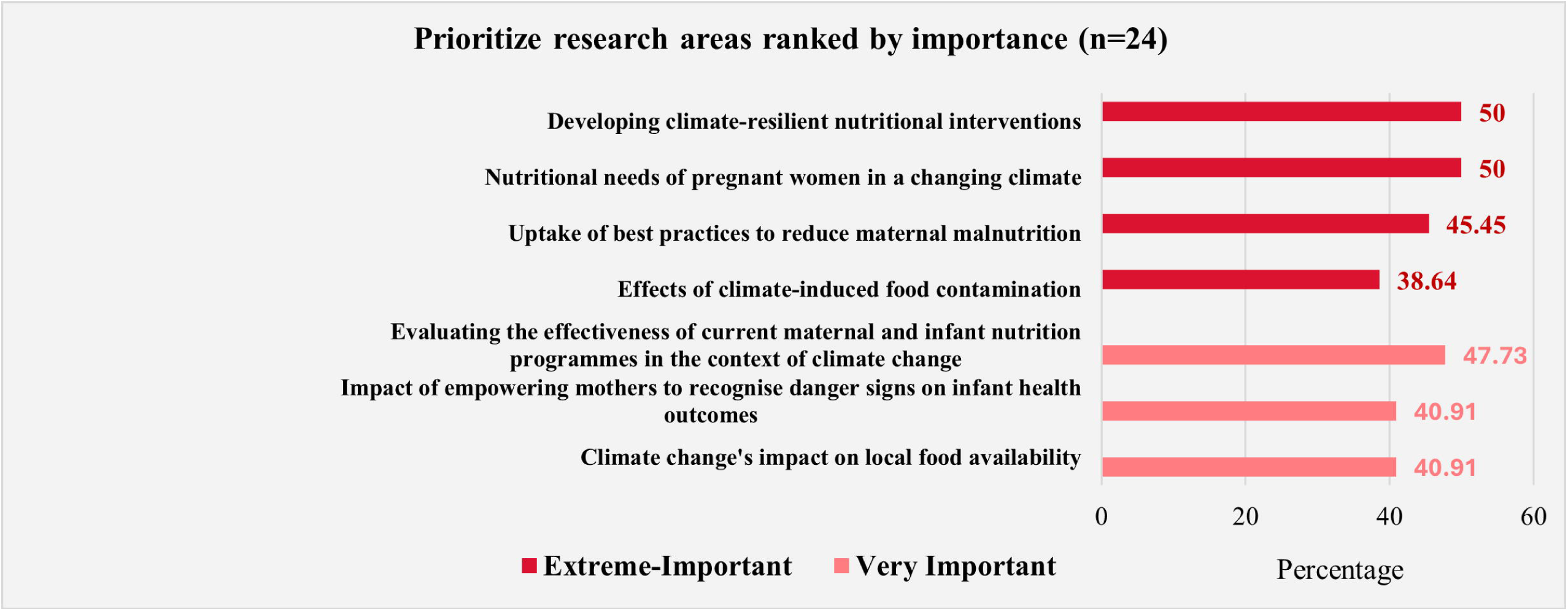
Key Research Areas ranked by importance in Maternal and Infant nutritional health impact by climate change.

## Qualitative Findings

### Characteristics of participants

Elven women and four men, including community members, health workers, and representatives from community-based organisations, participated in the session. Most of them represented the populations from local communities of the Matiari district; *see* Table 6

**Table 6.** Characteristics of Participants from the focus group.

| S# | Gender | Designation/Role | Affiliation/Experience |
| --- | --- | --- | --- |
| R1 | Female | Community Member | Community organization |
| R2 | Female | Community Midwife (CMW) | Community Health Volunteer |
| R3 | Female | Community Member | Local women's NGO |
| R4 | Male | Healthcare Provider | Community Welfare Organisation |
| R5 | Male | Farmer; Former health worker | Non-profit organization |
| R6 | Male | Social Worker | NGO Background |
| R7 | Female | Community Advocate | Awareness Sessions on Social Issues |
| R8 | Female | Lady Health Worker (LHW) | Maternal and child health |
| R9 | Female | Lady Health Supervisor (LHS) | Supervises LHWs |
| R10 | Female | Lady Health Worker (LHW) | Maternal and child health; vaccinations |
| R11 | Female | Community Midwife (CMW) | Health Training Background |
| R12 | Female | Community Midwife (CMW) | Private Clinic Services |
| R13 | Male | Social Worker | Community initiatives |
| R14 | Female | Research Associate | Maternal Health Projects |
| R15 | Female | Community Midwife (CMW) | Health Research Background |

### Identified themes

Several intersecting themes and sub-themes emerged from the data, illustrating how climate change interacts with gender, health, and social inequalities to impact maternal and infant nutrition and wellbeing. We organised themes into 4 major categories, including i) Climate disruption and health cascade effects, ii) Flood-induced crisis and maternal health vulnerabilities, iii) Gendered climate vulnerability and the triple Burden, and iv) Economic vulnerability and structural inequities.

## 1. Climate disruption and health cascade effects

Focus group participants and several survey respondents, in their qualitative comments, consistently described how shifting weather patterns, including delayed and extreme heat, unseasonal rainfall, and recurrent floods, had unexpected health effects and disrupted agricultural cycles, with serious consequences for families, particularly pregnant and breastfeeding women.

### 1.1 Shifting weather patterns and maternal health

Participants understood climate change not merely as extreme weather events such as heatwaves, floods, or heavy rain, but also as a gradual shift in seasonal patterns. These changes were perceived as a significant concern, producing unexpected and unforeseen health vulnerabilities for women and children. For instance, one participant pointed out both the absence of expected heat and the dangers of excessive temperatures:

As climate conditions have changed, usually in April it would be very hot, but now, because the temperature has not risen as before, things that used to happen no longer do. For example, mosquitoes die due to the heat, but now they are not dying because the heat isn’t as intense… the mosquitoes are more abundant. For instance, because the mosquitoes aren’t dying, malaria is spreading, and this disease is affecting children and mothers alike (Female participant).

Furthermore, participants described that this noticeable shift in weather patterns contributes to a range of adverse health outcomes, including heat stress, dehydration, fatigue, and pregnancy complications, mainly because families and communities are unprepared to cope with sudden environmental changes and their effects. These effects are compounded by physically demanding labour in extreme temperatures, which heightens the risks for expectant mothers. For example, a female participant said,

When the heat rises, whether she works at home or outside, she won’t stop working. She will continue working under all circumstances, and this can affect her blood pressure or cause other issues. If the heat is intense and affects the mother, it will obviously affect the child as well… (Female participant)

Another woman added, “*During unexpected heatwaves, many women continue working outdoors without adequate shade or water, which often results in dizziness and premature labour among pregnant women”.* In addition, several participants noted an increase in diseases, including diarrhoeal illnesses, skin infections, malaria, and dengue. For example, one male participant explained that *“stagnant water left behind after heavy rainfall or flooding becomes a breeding ground for mosquitoes. At the same time, delayed heat means mosquitoes are not dying as they once did”*. This change in weather patterns, therefore, creates conditions that increase malaria and dengue outbreaks, particularly affecting children and pregnant women.

Many participants and survey respondents highlighted that unexpected rainfall and flooding significantly disrupt access to essential maternal health services. Families are often unprepared for such events, leading to the closure of health facilities, especially in rural areas and the destruction of critical infrastructure, such as roads. These disruptions are especially concerning for antenatal and postnatal care. As one participant explained,

In heavy rain and flooding, women are mainly unable to access healthcare facilities, even in an emergency. Not only that, but during that time, there is no access to food or clean water, both of which are crucial for pregnant women (Female participant).

These accounts illustrate how disrupted weather patterns driven by climate change directly and indirectly undermine women’s health and well-being, increasing existing vulnerabilities within their communities.

### 1.2 Climate change and agriculture

Narrating their lived experiences with climate change, participants in both focus groups and the survey highlighted that climate change has severely disrupted agricultural cycles, resulting in lower crop yields and worsening nutritional outcomes for mothers and children. Some participants said shifts in seasonal patterns, such as delayed rainfall, unseasonal cold, and unexpected heat, are a “double burden” for both crops and health. As one participant explained:

Right now (April), the weather is cool, but it is not suitable for crops, as they need more heat to prepare for harvesting. After harvesting, it will get hotter; it is no use for crops, but many children will fall sick. And … heat strokes will be a more significant problem, meaning many children and adults, particularly pregnant women, can suffer from heat strokes and dehydration (Female participant).

Community participants observed that unpredictable weather has made traditional planting and harvesting schedules unreliable. For instance, a male focus group participant said, “*When rain arrives too late or too early, staple crops such as rice and wheat fail to mature properly, reducing household food supplies*.” Another added that “*similarly intense heat following the rainy season often damages vegetables and pulses, further shrinking dietary diversity*”.

There was a broad consensus among participants that these agricultural disruptions have direct consequences for maternal and infant nutrition. Several participants shared that food scarcity, combined with higher prices for vegetables and grains, forces families to reduce meal portions or skip protein-rich foods. A female participant said that *“pregnant women are eating less or not eating proper nutritious food because it has become expensive, and crops keep failing due to changing weather.”* Focus group participants, especially lady health workers and community midwives, were of the view that such conditions increase the risk of anaemia, low birth weight, and other pregnancy-related complications.

## 2. Flood-induced crisis and maternal health vulnerabilities

Participants recalled Pakistan’s 2022 mega floods as a convergence of crises, including displacement, income loss, water contamination, and health service disruption, that particularly harmed maternal and child health. The floods created vulnerabilities that persisted well beyond the immediate emergency and were reiterated by many participants:

When the flood came, all our crops were destroyed. Houses were also destroyed, and we didn’t receive the wages that were due for our work. And if there are pregnant women, we mostly meet them in the evenings or at night because they also do outdoor work, household work, and they have children too. And their nutritional needs were not met at all (Male participants).

Community members explained that it was not only the flood but also prolonged moisture and stagnant water that had a greater impact; for instance, the mosquito population increased, thereby increasing malaria risk. Some participants’ survey comments suggested that flooding also disrupted the food supply chain, leading to food shortages and malnutrition, which have been particularly harmful for pregnant women and infants who need proper nutrition for healthy development. A community midwife observed:

Because of climate change, the floods and other factors have had a particularly bad impact on our area. When there were displacements and migrations, when people’s homes were destroyed, when there was nothing for people to eat, these negative impacts affected all of us, especially pregnant and breastfeeding women. Mothers, children, and adolescent girls were affected more because there was a lack of food, no proper shelter for displaced families, and due to stagnant flood water, there were more diseases. (Female participant)

Some participants reported that, due to flooding, they lost crops, resulting in a loss of both food and income and limiting families’ ability to afford nutritious diets essential for maternal and foetal health. A participant, who was a social worker, recalled:

The loss of income from heavy rain and flooding meant our food and crops were destroyed, so we didn’t have any income. So, of course, we could not buy the food, the nutritious food, so pregnant women were impacted. And another very important thing is that there were premature births because pregnant women were stressed during the flooding. And there were no safe roads or any paths to access the healthcare facilities. So, women had deliveries in their homes, maybe at seven months, premature babies. They should have been in a hospital, but they couldn’t get to one. So, babies were very low birth weight; some of them weighed only 1 kilogram (Male participants).

These participants’ accounts illustrate how repeated flooding creates multiple issues, increasing vulnerabilities that lead to a crisis impacting their livelihood, health, and well-being.

## 3. Gendered climate vulnerability and the triple burden

### 3.1 Working hard to survive

From participants’ perspectives, both men and women labour intensively to survive under the compounded pressures of poor socioeconomic conditions, which are partly exacerbated by climate change. Families often work collectively to meet basic nutritional needs, as one male participant explained, *“to have food on the table/eat something.”* However, participants emphasised that women, including those who are pregnant, often bear a disproportionate burden during climate extremes due to their overlapping roles as agricultural workers, caregivers, and homemakers. This “triple burden” is frequently intensified during crises yet remains invisible, mainly in broader climate discussions. To illustrate this, a lady health worker described women’s relentless daily schedule:

Their daily routine starts with the Fajr (dawn) prayer; they milk buffaloes or cows, prepare breakfast, feed the children, do the children’s chores, clean the house, and then go to the fields. Staying continuously standing in the field also causes health issues for them. They go to work in the fields with the mindset that they have to earn the day’s wage no matter what… Then they also cut a lot of grass from the fields, and they have to carry the bundle home… Then, in the afternoon, they come home to cook for their family members, like mother-in-law, father-in-law, and children, and after that, they go back to the fields. In the rush of work, their own nutritional needs are neglected” (Female participant).

Further emphasising the risks during pregnancy, another lady health worker explained:

For pregnant women, I would say that now that it is the wheat harvesting season, every pregnant woman, whether 40 days into pregnancy or 3 or 4 months pregnant, is focused on cutting or gathering wheat so they can eat it later. They are not going for their check-ups, they are not getting their tests done, they are not going to the hospital because these three or four days are the wheat season… Out of five women, three are fine, but we keep the others on a course of (name of medicine) … Because they do not go for check-ups, they are at risk of abortion (Female participant).

Consequently, women’s routines, beginning before dawn and ending late at night, combine agricultural labour, household chores, childcare, and caregiving for older adults, leaving little opportunity for rest or self-care. Some participants noted that male family members provide limited support, particularly when they are working long hours as breadwinners. Others contested this, highlighting that support varies across households. Nonetheless, the unrelenting workload, combined with environmental stressors, contributes to chronic fatigue, mood swings, and emotional burnout.

Participants also highlighted the absence of mental health services and awareness. Mental health issues, including stress, depression, and anxiety, often go unrecognised or are misdiagnosed. Access to medication and follow-up care is limited, while social stigma further prevents women from seeking support. In this context, the physical and mental burdens borne by women emerge as a critical, yet underacknowledged, consequence of climate-induced socioeconomic pressures. Participants connected women’s unrelenting workload to broader climate-induced crises, highlighting how extreme events increase existing vulnerabilities. These accounts underscore that gendered vulnerabilities are intensified when environmental shocks intersect with the physical, nutritional, and mental strains women already endure, making them particularly vulnerable during climate disasters.

### 3.2 Lack of social welfare

Participants emphasised the hardships women endure to support their families, noting the absence of adequate national welfare support as a critical gap. In particular, maternity leave and financial support for pregnant women in informal or agricultural work remain largely unavailable. Families’ health and survival often depend on the labour of both women and men; without alternative support during pregnancy or postpartum periods, women’s absence from work can generate additional financial and emotional stress for the entire household, while compromising maternal well-being. A local researcher working on maternal and adolescent reproductive health highlighted the limitations of existing programmes:

We have heard there are social welfare programmes like Benazir Income (Pakistan’s national social protection programme, Benazir Income Support Programme (BISP) and Mumta (Mother and Child Support Programme (MCSP), but they don’t support all women, and women face many challenges from registration to obtaining cash or other support through such programmes. There are many reasons, but the major one is that men run most of these programmes from top to bottom, so it is challenging, even culturally restricted, for many women even to go and ask for help from men (Female participants).

Participants discussed examples of women who were unable to access cash transfers during critical periods, forcing them to continue physically demanding work while pregnant or caring for newborns. A participant shared, in one case, *“a woman skipped her postnatal check-ups because she had to continue harvesting crops to feed her family, as no financial support was available to replace her labour”*.

This evidence shows that the lack of social welfare programmes exacerbates the physical, emotional, and financial burdens women already face, reinforcing cycles of vulnerability in contexts where climate pressures, poor nutrition, and limited healthcare intersect.

## 3. Economic Vulnerability and Structural Inequities

### 4.1 Poverty and climate vulnerability

Participants highlighted that poverty functions both as a cause and a consequence of climate vulnerability, creating a difficult-to-break cycle. Many families depend on subsistence agriculture yet often produce crops they cannot afford to consume themselves. Instead, they are compelled to sell the harvest to meet immediate cash needs, only to repurchase the same food at higher prices, leaving little room for savings or dietary diversity. As one participant observed, “*It is expensive. Even those who produce it can’t afford to consume it… They have to give it to the market, get money, and then buy the same food again*” (Male participant).

Most participants view climate-related disruptions as further increasing this cycle of dependence on unstable markets and low income. For instance, a female participant said, *“Floods, heatwaves, and heavy rainfall destroy crops, reduce crop yields, and erode household assets, thereby reinforcing poverty and increasing malnutrition, particularly among women and children”*. The financial precarity limits access to healthcare, nutritious food, and other essential services, making families more vulnerable to both immediate shocks and long-term health consequences.

Several participants emphasised that poverty underpins many of these challenges. One participant explained:

Suppose someone earns 500 rupees a day, what can they manage with that? What can they cover with 500 rupees? If there are five people in the household, how can they manage with so little money? So, the main issue is poverty” (Female participant).

Participants emphasised that poverty not only constrains families’ ability to adapt to changing environmental conditions but also increases the health and nutritional risks associated with climate stressors. Participants’ accounts reveal that addressing climate vulnerability requires tackling these deep-rooted economic inequalities alongside environmental interventions.

### 4.2. Global health inequality and unfair debt systems

Within the context of global climate change, participants drew a direct connection between Pakistan’s external debt and the state’s limited capacity to provide public relief. The burden of debt was seen not only as an economic constraint but also as a structural barrier to climate resilience and health equity. For example, a male participant demanded that *“Foreign governments, especially Western, should write off Pakistan’s debt instead of giving Pakistan peanuts in charity.”* This sentiment reflects a broader frustration with the global financial systems, which participants perceived as perpetuating dependency rather than enabling recovery. A male participant emphasised the role of international diplomacy, suggesting: *“The UK could press the International Monetary Fund (IMF) to cancel Pakistan’s debt.”*

Participants linked these macroeconomic constraints to everyday struggles, particularly food insecurity. They described how reduced crop yields due to climate shocks, such as floods and heatwaves, were compounded by systemic poverty and rising costs, making food inaccessible for many rural households. For example, one woman explained, *“Our family had to skip meals after the floods destroyed our crops and livestock, while prices of basic staples doubled in local markets”*.

Most participants saw poverty as the most significant issue and believed the government could provide relief if international institutions wrote off the country’s debt.

## Discussion

This study represents a unique insight into the lived experience of those working in maternal and infant health in Sindh, Pakistan. The participants represented those in front-line services (micro system), in academic practices and research (meso system) and policy development and strategy (macro system). The majority (78 of 88) expressed extreme or very high concern that nutritional awareness was a priority risk for pregnant women, alongside access to nutritional support. The impact of low nutritional awareness may be linked to other priority concerns for women in pregnancy, including dehydration, weakness and exhaustion, and limited access to foods linked to environmental factors and climate change. Participants recognised that in their front-line practice and based on their research knowledge, evidence indicates that heat-related infant mortality and low birth weight are significantly increasing [11]. Previous studies emphasise the need for further research on the effects of heat exposure on infant health, indicating that extreme temperatures can lead to adverse outcomes, including low birth weight and even death [25].

The prioritisation produced in this survey and during workshops suggested that the shifting weather patterns had a detrimental effect on agricultural activities. Unseasonal rainfall and recurrent floods are affecting household incomes, and poor yields and crop failures are increasing the precarity of whole communities. The gradual shift in climate was deemed more significant than the environmental ‘shocks’ the flooding experienced in recurring years. Pest control and access to drinking water are constant challenges. These instabilities and the wide range of anxiety-provoking climate impacts are understudied and are challenging to quantify at the local level [26]. This is because the localised experience of household communities is missing from current evaluations of the climate risks to lives and livelihoods. Figure 3 below outlines micro, meso and macro factors that intersect and influence communities’ health and wellbeing in changing climatic conditions of Pakistan.

**Figure 3.**
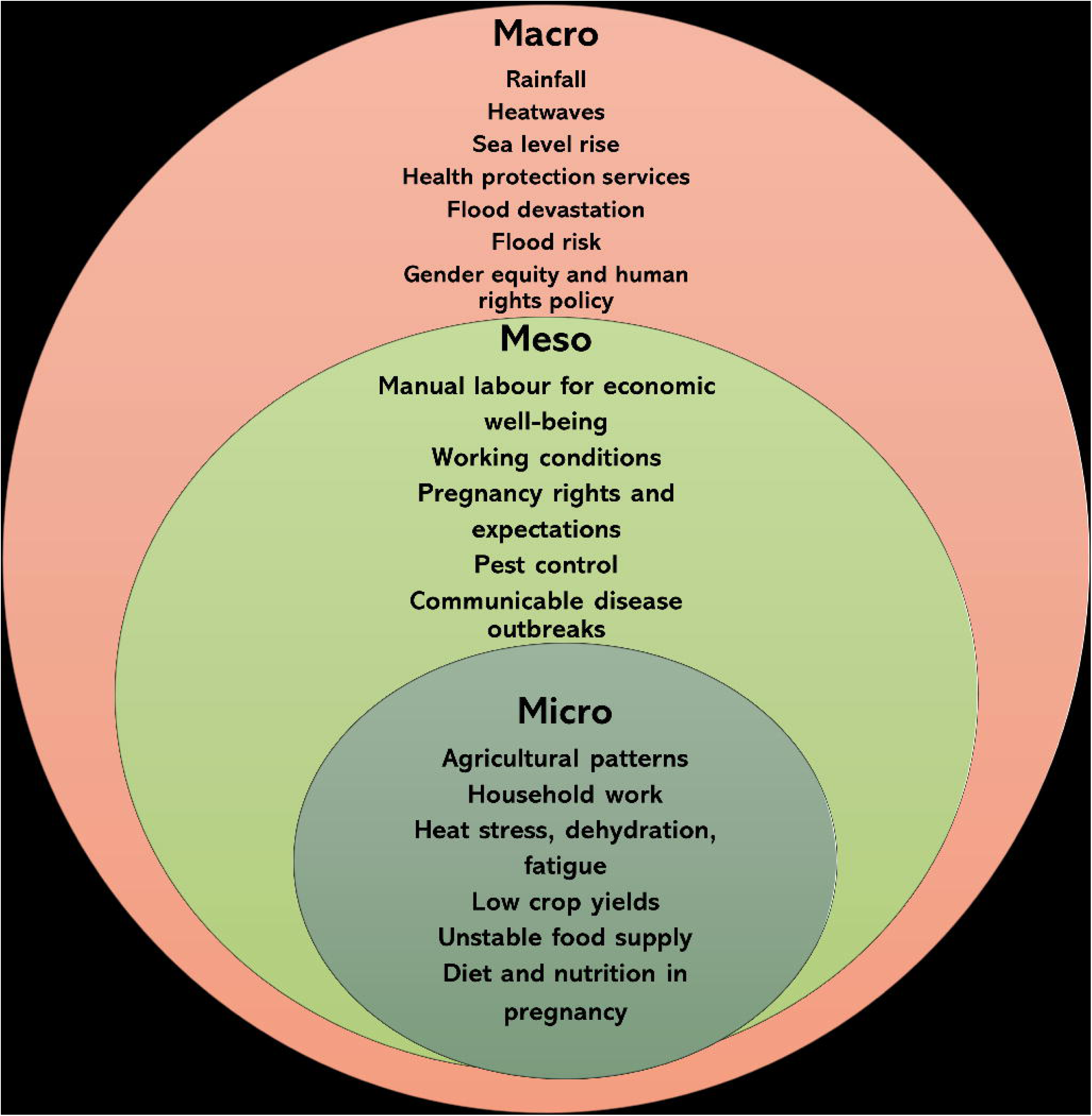
Intersecting Micro-Meso-Macro factors.

### Macro perspectives

The poverty and precarity at the household level are well recognised and long-standing macroeconomic risks acknowledged internationally. Low yields have a ‘multiplier effect’ on the cost of living, and it is thought that 60% of Pakistan’s population relies on agriculture [27]. The threats to long-term food security remain a problem nationally, due to reduced agricultural productivity [28]. Agricultural communities are disproportionately affected, and rural populations experience entrenched social and health inequities. This is where access to healthcare and nutrition is limited [7].

Developing countries owe huge debts that grow twice as fast as those of rich countries, with countries like Malawi having 71.2% poverty rates because debt payments divert funds from health services. However, countries that maintained strong local agricultural communities did better at reducing poverty than those focused on mining or large plantations, suggesting that solutions to climate-health problems should strengthen community knowledge, local food systems, and traditional ways of caring for pregnant women and infants [29–31].

Challenges are compounded by structural inequities, such as gender roles, economic precarity, and social exclusion, which tend to restrict women’s ability to rest, access care, or make autonomous health decisions [32, 33]. Gender equality is a strategic ambition to alleviate poverty in Pakistan, but the lack of progress is increasing the vulnerability of poor communities [34]. This is evident in the disproportionate impact of climate change on maternal and infant nutrition, highlighting how environmental changes exacerbate malnutrition and health disparities [35] and in the critical relationship between maternal nutritional status and child growth outcomes, indicating that poor maternal health is significantly associated with stunting and wasting in children in Pakistan [36]. These negative health outcomes necessitate further research into mitigation and adaptation strategies that include social justice, community interventions, regional governance and policy to reduce gender inequality. These would underpin the implementation of locally acceptable methods to protect pregnant women in their communities.

### Meso perceptions

Climate change continues to undermine social determinants of health and disproportionately affect the most vulnerable populations, including women and infants in poor communities. Social protection plays a key role in countering the impacts of climate change at a regional level. However, many are least prepared for the climate crisis they face; for example, 91.3% of people in the 20 most climate-vulnerable countries lack any form of social protection coverage [1,37].

Our findings reveal that a lack of awareness regarding proper nutrition, crop damage and water contamination remains a critical barrier, a factor noted in prior studies [9,17,38]. These findings suggest that, at a microsystems level, household poverty and health literacy urgently need to be addressed to enable young women to receive further guidance on how they live and work in their communities during pregnancy. The existing literature consistently highlights the compounded health risks posed by climate change and socioeconomic constraints, particularly for adverse pregnancy outcomes and mothers’ mental health in extreme heat and floods [39,40]. Aziz and Anjum emphasise the need to integrate cultural and gender perspectives into climate adaptation strategies, which is crucial for addressing the specific vulnerabilities of women in rural contexts [41]. This aligns with the broader policy discourse at the national and regional scales on the accessibility and availability of health services during climate disasters.

### Micro perspectives

Further, the findings show that participants are aware of the risks associated with extreme heat and floods significantly impact women’s physical and mental well-being. Challenges related to pregnancy preparedness, caring for newborns, and hard agricultural work tend to exacerbate health risks for individuals. Failing to connect community health awareness and the health system with climate action results is a missed opportunity for maternal and infant health outcomes [42]. This knowledge gap among healthcare providers underscores the need to improve education and training for healthcare professionals to mitigate the impact of climate change on maternal health [43].

### Existing local interventions

There are social protection programmes, including the Benazir Income Support Programme (BISP) and the Sindh Social Protection Agency; however, these are neither fair nor accessible to many vulnerable women [44–46]. Thus, strengthening these initiatives can reduce health inequalities by ensuring that maternal and infant health services reach those most disadvantaged. In Pakistan, recent evidence from Sindh and similar contexts supports this. For example, a cluster-randomised controlled trial in Dadu District showed that larger cash transfers reduced wasting at 6 months, and that all cash and voucher-based intervention modalities improved height-based growth at both 6 months and 1 year among children under five and their mothers [47]. The Pakistan Maternal Nutrition Strategy 2022-2027 reports [48] that only about 28% of women of reproductive age meet the minimum dietary diversity, with rates even lower in rural and low-wealth groups, highlighting how nutritional inequities can be addressed through targeted welfare interventions [49]. Evaluations of the Sehat Sahulat Program indicate high patient satisfaction and reduced financial burden for low-income households, though they also highlight administrative and awareness challenges that constrain equitable access [50]. The “Enough Food Model” and other food security initiatives similarly show promise in improving maternal nutrition among food-insecure and low-resource populations [51]. These studies suggest that equitable welfare interventions, including cash transfers, free or subsidised healthcare, nutrition-focused programmes, and support for food security, combined with strong administrative design, monitoring, and awareness, are key to narrowing maternal/infant health inequalities in settings like Sindh.

Our study highlights the potential to develop more effective and sustainable targeted interventions to strengthen health systems capable of addressing the multifaceted challenges of climate change. Climate change undermines many of the social determinants for good health, such as livelihoods, equality and access to health care and social support structures, with these climate-sensitive health risks disproportionately felt by the most vulnerable and disadvantaged, including women, children, poor communities, migrants or displaced persons, and those with underlying health conditions (malnutrition) [52]. It emphasises the critical need to bridge knowledge gaps, foster robust community engagement, and incorporate cultural considerations into health interventions.

### Strengths and Limitations

This study provides valuable insights into local knowledge and highlights the complexity of maintaining nutritional well-being during pregnancy in an agricultural community facing climate challenges. It includes diverse stakeholders, but the sample may not fully capture all perspectives, particularly those of marginalised or highly remote communities. Additionally, the study’s findings are region-specific, and their generalisability to other areas affected by climate change should be approached with caution. Methodological constraints, such as reliance on self-reported data in surveys and workshops, may introduce biases, though efforts were made to triangulate findings with qualitative insights.

### Recommendations

Our findings on climate-health challenges, particularly the lack of nutritional awareness and maternal dehydration identified by participants, cannot be separated from more profound historical injustices that still shape health outcomes today.

Future research should explore the long-term impacts of climate-induced challenges on maternal and infant health, particularly as climate variability intensifies. There is also a pressing need to develop and test interventions that address the intersecting issues of nutrition, maternal and infant health, and healthcare access in the context of climate resilience. Moreover, studies should aim to investigate the effectiveness of community-driven approaches and culturally tailored strategies in mitigating these risks. Addressing gaps in including vulnerable populations in research and policy planning will be essential for developing comprehensive solutions.

## Conclusion

The thematic categories identified in this research suggest a comprehensive, intersectional approach that is required for effective climate-resilient social welfare systems. These are needed to address multiple, interconnected vulnerabilities associated with climate change and maternal and infant nutrition. Actions to manage health risks are required at multiple levels to ensure that climate action does not deepen existing inequalities.

The engagement and participation demonstrated in this research in Sindh, Pakistan, are essential to ensure that interventions are planned with eco-social approaches. This is more likely to result in interventions that reflect local community priorities. Nutritional improvements for pregnant women are critically important in the context of a changing climate and in an agrarian economy. More marginalised communities require health systems that are sensitive to the intersectional challenges and co-produced interventions that reflect the realities of their daily lives.

## Data Availability

The corresponding authors will review requests for data access to ensure that the use of the data is in accordance with the terms of ethics approvals and principles. Upon reasonable request, access to the data will be made available following publication.

## Supporting information

“For the purpose of open access, the authors have applied a Creative Commons Attribution (CC BY) licence to any Author Accepted Manuscript version of this paper, arising from this submission.”

## Authors’ contributions

**Conceptualisation**: Sadiq Bhanbhro and Zahid Memon.

**Formal analysis**: Sadiq Bhanbhro, Zahid Memon, Wardah Ahmed, Fizza Ansar

**Methodology**: Sadiq Bhanbhro, Hora Soltani

**Project administration**: Sadiq Bhanbhro, Zahid Memon, Wardah Ahmed, Fizza Ansar

**Writing original draft:** Sadiq Bhanbhro, Wardah Ahmed, Fizza Ansar

**Writing** – **review & editing:** Sadiq Bhanbhro, Zahid Memon, Wardah Ahmed, Fizza Ansar, Gulfishan Tahir, Nadia Agha, Sally-Fowler Davis, Hora Soltani

## Supporting information

S1 Table GRAMMS research checklist.

S2 Table Participants’ demographic information

S1 Figure 1. Awareness and Concern about Maternal & Infant Nutritional Health due to Climate Change

S2 Figure 2. Key Research Areas ranked by importance in Maternal and Infant nutritional health impact by climate change

S3 Figure 3. Intersecting Micro-Meso-Macro factors S1 Survey questionnaire

